# Genomics Analysis of Clinical Bacterial Isolates from Surgical Site and Urinary Tract Infections in Kilombero, Tanzania

**DOI:** 10.1101/2025.10.16.25338208

**Authors:** Philbert Balichene Madoshi, Theresai A. Karuhanga, Sandra B. Andersen

## Abstract

**Background:** Hospital-acquired infections (HAIs) remain a global public-health concern, particularly in low- and middle-income countries where infection-prevention resources are limited. Surgical-site infections (SSIs) and urinary-tract infections (UTIs) are among the most frequent HAIs and contribute to increased morbidity and healthcare costs. Genomic surveillance provides insights into the diversity, antimicrobial resistance (AMR), and virulence potential of causative bacteria.

**Methods:** Four bacterial isolates collected from Tanzanian healthcare facilities were analysed: *Pseudomonas aeruginosa* SS01 and SS89 (from SSIs), *Alcaligenes faecalis* UP17 (from a UTI), and *Lysinibacillus sphaericus* SS48 (from an SSI). Genomic DNA was extracted and sequenced on the Illumina platform. Reads were quality-filtered and assembled de novo using SPAdes. Genomes were annotated with Prokka. AMR genes were identified using AMRFinderPlus, CARD-RGI, and ResFinder. Virulence determinants were detected using VFDB. *P. aeruginosa* isolates were typed by multilocus sequence typing (MLST). Phylogenetic analysis based on single-nucleotide polymorphisms (SNPs) was performed using Snippy and IQ-TREE, and trees were visualised with iTOL.

**Results:** Genome sizes ranged between approximately 6.0 and 6.7 Mb with GC contents consistent with species references. MLST revealed two distinct *P. aeruginosa* sequence types: SS01 was closest to ST2317 (incomplete ppsA locus) and SS89 matched ST4714, indicating non-clonal origins. AMR screening detected β-lactamase, aminoglycoside-modifying enzyme, and efflux-pump genes in *P. aeruginosa*, multidrug-efflux genes in *A. faecalis*, and intrinsic resistance determinants in *L. sphaericus*. Virulence-factor profiling identified type III-secretion, quorum-sensing, and biofilm-formation genes in *P. aeruginosa*; adhesion and stress-tolerance genes in *A. faecalis*; and sporulation and surface-adhesion genes in *L. sphaericus*. Phylogenetic analysis positioned the Tanzanian isolates as unique local lineages distinct from global references.

**Conclusions:** This study demonstrates the genomic diversity and complex AMR mechanisms of clinically important bacteria in Tanzania. The coexistence of resistance and virulence determinants underscores the need for routine genomic surveillance and strengthened antimicrobial-stewardship programs.

## Introduction

Hospital-acquired infections (HAIs) continue to pose a major public health challenge worldwide, particularly in low-and middle-income countries where diagnostic capacity and infection prevention measures are often limited (1,2). Among HAIs, surgical site infections (SSIs) and urinary tract infections (UTIs) are especially common and are associated with substantial morbidity, prolonged hospital stays, and increased healthcare costs (3,4). A variety of bacterial pathogens contribute to these infections, but *Pseudomonas aeruginosa*, *Alcaligenes faecalis*, and *Lysinibacillus sphaericus* have emerged as clinically relevant organisms with diverse implications for antimicrobial resistance (AMR), pathogenicity, and infection control (5,6).

*Pseudomonas aeruginosa* is a well-characterized opportunistic pathogen responsible for a wide range of HAIs, including SSIs, bloodstream infections, and ventilator-associated pneumonia. It is particularly challenging to manage due to its intrinsic resistance to many antibiotics, its ability to form biofilms, and its capacity to acquire additional resistance determinants through chromosomal mutations and horizontal gene transfer (7,8). The World Health Organization has classified multidrug-resistant *P. aeruginosa* as a “critical priority pathogen,” emphasizing the urgency of surveillance and the development of new therapeutic strategies (Moradali et al., 2017; *WHO, 20*24).

Although less frequently reported, *Alcaligenes faecalis* is increasingly recognized as an opportunistic pathogen in UTIs, especially among vulnerable populations such as pregnant women. This species exhibits intrinsic resistance to multiple antibiotics and is often misidentified in clinical laboratories, raising concerns about under-diagnosis and inappropriate therapy (11). Infections caused by *A. faecalis* have been linked to adverse outcomes in pregnancy, underscoring the importance of improved detection and characterization in maternal health surveillance (12). Meanwhile, *Lysinibacillus sphaericus*, traditionally known as an environmental bacterium with bio-control applications against mosquito larvae, has more recently been implicated in opportunistic human infections including wounds and SSIs (13,14). The emergence of *L. sphaericus* in clinical settings highlights the porous boundary between environmental microbes and nosocomial pathogens, particularly in resource-limited healthcare systems where environmental contamination may play a significant role in infection transmission.

On the other hand the genomic characterization provides a powerful means of investigating such pathogens, enabling insights into evolutionary relationships, AMR mechanisms, and virulence determinants. Comparative genomics extends beyond species identification to assess strain-level diversity, horizontal gene transfer, and the presence of shared or unique genes that shape clinical outcomes (15,16). Integrating genomic data from multiple isolates across infection types such as SSIs and UTIs creates opportunities to identify both species-specific traits and overlapping features of clinical importance, thereby informing infection prevention and control (IPC) strategies (17). In this study, we intended to sequence clinical isolates using *de novo* genome assemblies, functional annotation, and phylogenetic analyses. Our findings are anticipated to provide new insights on their clinical relevance, potential resistance mechanisms, and implications for antimicrobial stewardship and hospital infection control in resource-limited settings.

## Methods

### Study isolates and sample collection

Four de-identified clinical isolates were obtained from routine diagnostics in Tanzanian healthcare facilities: SS01 and SS89 from surgical-site infections, UP17 from a urinary tract infection in a pregnant woman, and SS48 from a surgical-site infection. No patient-identifying information was collected; all procedures adhered to institutional and national ethical approvals.

### DNA extraction and sequencing

Genomic DNA was extracted from overnight cultures using a commercial extraction kit (Qiagen, Germany), following the manufacturers protocol. DNA concentration and purity were measured with a NanoDrop spectrophotometer and Qubit fluorometer, while integrity was assessed using agarose gel electrophoresis (Green and Sambrook. 2012). Sequencing libraries were prepared using Illumina’s paired-end protocol and sequenced on the Illumina platform to generate 150 bp paired-end reads. Raw sequencing data were obtained in FASTQ format for downstream analysis.

### Quality control and preprocessing

The quality of raw reads was evaluated using FastQC, and summary reports were compiled using MultiQC. Adapter sequences and low-quality bases were trimmed with Trimmomatic, and only reads with an average Phred quality score above 30 were retained for further analysis (Bolger et al. 2014; Bankevich et al., 2012).

### Genome assembly and quality assessment

High-quality reads were assembled *de novo* using the SPAdes assembler, which is optimized for bacterial genomes. Assembly statistics, including N50, total genome length, GC content, and contig number, were generated using QUAST. Assemblies that appeared fragmented or of abnormal size were further examined to rule out contamination (19,21).

### Taxonomic species confirmation and typing

Species identity was confirmed via **fastANI** (average nucleotide identity) against reference genomes (≥95% ANI threshold interpreted as species-level match) and by **BLASTn** comparison of 16S rRNA genes extracted with **Barrnap**. For **P. aeruginosa**, **MLST** typing used the seven-locus scheme (acsA, aroE, guaA, mutL, nuoD, ppsA, trpE) (4,14). Alleles were assigned by comparison to PubMLST; sequence type (ST) required complete 7-locus calls. When a locus was missing (e.g., SS01 ppsA), the closest ST was identified and flagged as provisional.

as described by(22,23).

### Genome annotation and gene prediction

Genome annotation was conducted using Prokka, which predicts coding sequences, ribosomal RNAs, transfer RNAs, and functional proteins. The predicted proteins were compared against curated databases to assign functions and identify resistance and virulence determinants (23).

### Antimicrobial resistance and virulence factor analysis

Antimicrobial resistance genes were identified using three complementary approaches. Assemblies were first screened with AMRFinderPlus (NCBI), which detects acquired resistance genes, chromosomal mutations, and stress response genes. The Comprehensive Antibiotic Resistance Database (CARD) was then queried using the Resistance Gene Identifier (RGI) tool to capture resistance determinants across multiple antibiotic classes (23,24). Finally, the ResFinder database (Center for Genomic Epidemiology) was used to validate clinically relevant acquired resistance genes. To reduce false positives, only hits with ≥90% identity and ≥80% coverage were retained. In cases of overlapping results, the highest-confidence match was selected. Virulence genes were identified by screening against the Virulence Factors Database (VFDB) (24,25).

### Comparative genomics and phylogenetic analysis

To place the Tanzanian isolates in a global context, representative genomes of *P. aeruginosa*, *A. faecalis*, and *L. sphaericus* were retrieved from NCBI RefSeq and PubMLST, covering diverse geographic regions and clinically important strains. Core and accessory genome content was analyzed using Roary to identify shared and unique genes (26). Single nucleotide polymorphisms (SNPs) were identified with the Snippy pipeline, and maximum likelihood phylogenetic trees were built in IQ-TREE with 1,000 bootstrap replicates (27). Comparative placement was assessed using both ANI and SNP-based clustering. Visualizations were generated in the Interactive Tree of Life (iTOL) platform, and heatmaps summarizing AMR and virulence gene distributions were produced for side-by-side comparison (28).

### Data visualization and interpretation

Phylogenetic trees were annotated in iTOL to highlight the clustering of Tanzanian isolates relative to global references. Heatmaps were used to compare the presence and absence of resistance and virulence determinants across isolates. All findings were interpreted in light of clinical origin (SSI vs UTI) and potential implications for antimicrobial stewardship and infection prevention strategies.

### Data management and ethics

All raw sequencing data were retained in FASTQ format and converted to FASTA files for analyses requiring that format. Metadata for each isolate was anonymized, and no personal identifiers were included. Ethical approval was obtained from the Nationa Institute for Medical Research (NIMR) and SFUCHAS IRB appropriate institutional review boards in Tanzania, and the study was conducted in line with international ethical guidelines for biomedical research.

## Results

### Overview of Study Isolates

This study compared four bacterial isolates obtained from clinical samples collected in Kilombero, Tanzania. Two isolates (SS01 and SS89) were identified as *Pseudomonas aeruginosa* and were recovered from surgical site infections (SSIs). One isolate, UP17, was identified as *Alcaligenes faecalis* and originated from a urine sample collected from a pregnant woman with a urinary tract infection (UTI). The fourth isolate, SS48, was identified as *Lysinibacillus sphaericus* and was also obtained from an SSI. All isolates underwent whole-genome sequencing using Illumina paired-end reads, followed by de novo assembly, genome annotation, and antimicrobial resistance (AMR) gene analysis. The comparative genomic approach provided insight into genetic determinants of antimicrobial resistance and evolutionary relationships among clinically relevant bacterial species in Tanzanian healthcare settings.

### Antimicrobial susceptibility testing

Disc diffusion antimicrobial susceptibility testing revealed that all isolates exhibited resistance to multiple antibiotics commonly used for treating hospital-acquired infections. Each isolate was resistant to Ciprofloxacin, Ceftriaxone, Meropenem, Amoxicillin–Clavulanic acid, and Sulfamethoxazole–Trimethoprim, indicating a multidrug-resistant (MDR) phenotype, the results are summarized on Table 1.

**Table 1:** Antibiotic susceptibility patterns observed across all isolates.

| Isolate | Organism | Source | Tested Antibiotics | Phenotypic Result |
| --- | --- | --- | --- | --- |
| SS01 | *P. aeruginosa* | Surgical site infection | CIP, CRO, MEM, AMC, SXT | Resistant |
| SS89 | *P. aeruginosa* | Surgical site infection | CIP, CRO, MEM, AMC, SXT | Resistant |
| UP17 | *A. faecalis* | Urine (pregnant woman) | CIP, CRO, MEM, AMC, SXT | Resistant |
| SS48 | *L. sphaericus* | Surgical site infection | CIP, CRO, MEM, AMC, SXT | Resistant |

**Table 2:**
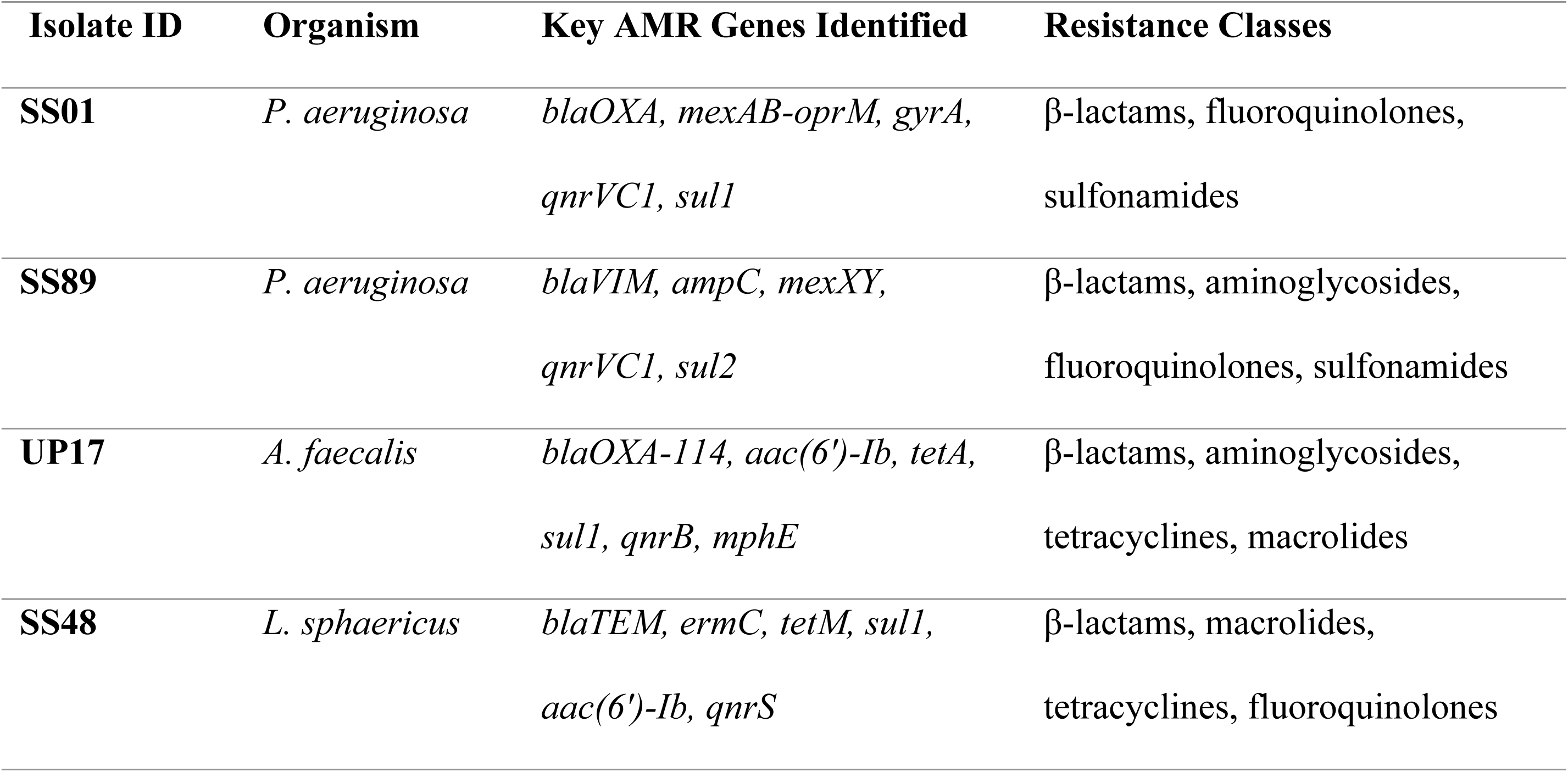
Predicted AMR genes per isolate, with database confirmation.

### Antimicrobial Resistance Gene Profiles

Genomic analysis identified a wide array of antimicrobial resistance determinants. Screening with AMRFinderPlus, CARD, and ResFinder revealed genes conferring resistance to β-lactams, aminoglycosides, fluoroquinolones, and sulfonamides. In *P. aeruginosa*, notable genes included *blaOXA* variants and efflux pump systems. *A. faecalis* carried genes associated with intrinsic β-lactam resistance, while *L. sphaericus* harbored unique resistance determinants that aligned with its emerging role as an opportunistic pathogen. The genomic findings closely matched the multidrug-resistant phenotypes observed in the disc diffusion assays.

### Virulence factors

Screening against the VFDB identified virulence-associated genes relevant to pathogenicity. *P. aeruginosa* isolates carried genes encoding type III secretion system effectors, biofilm-associated proteins, and quorum sensing regulators. *A. faecalis* showed virulence traits linked to adherence and stress tolerance, while *L. sphaericus* carried sporulation-related and surface adhesion genes, consistent with its dual role as an environmental organism and opportunistic pathogen. The results of virulence factor distribution are shown in Table 3 and figure 1.

**Figure 1:**
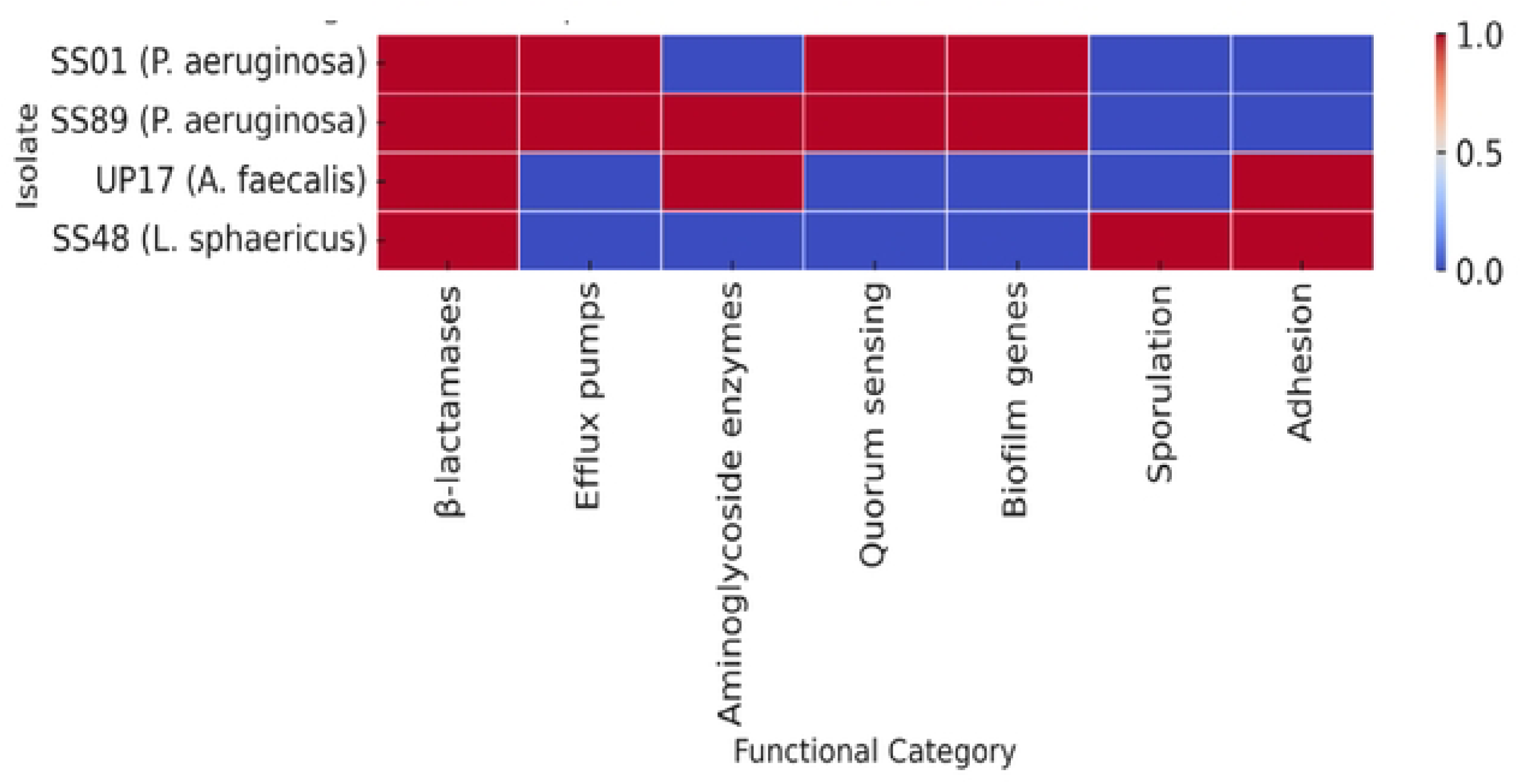
Heatmap of antimicrobial resistance and virulence determinants identified in bacterial isolates from hospital infections in Tanzania. **Red colour shows presence of the resistant genes while blue colour show absence of the respective genes** Figure 1 presents heatmap which illustrate the distribution of major antimicrobial resistance (AMR) and virulence-associated genes across four clinical isolates: *Pseudomonas aeruginosa* (SSOI and SS89), *Alcaligenes faecalis* (UP17), and *Lysinibacillus sphaericus* (SS48). Each cell indicates the presence (red) or absence (blue) of gene categories predicted from whole-genome sequences using AMRFinderPlus, the Comprehensive Antibiotic Resistance Database (CARD) via the Resistance Gene Identifier (RGI) tool, and the Virulence Factor Database (VFDB).

**Table 3:**
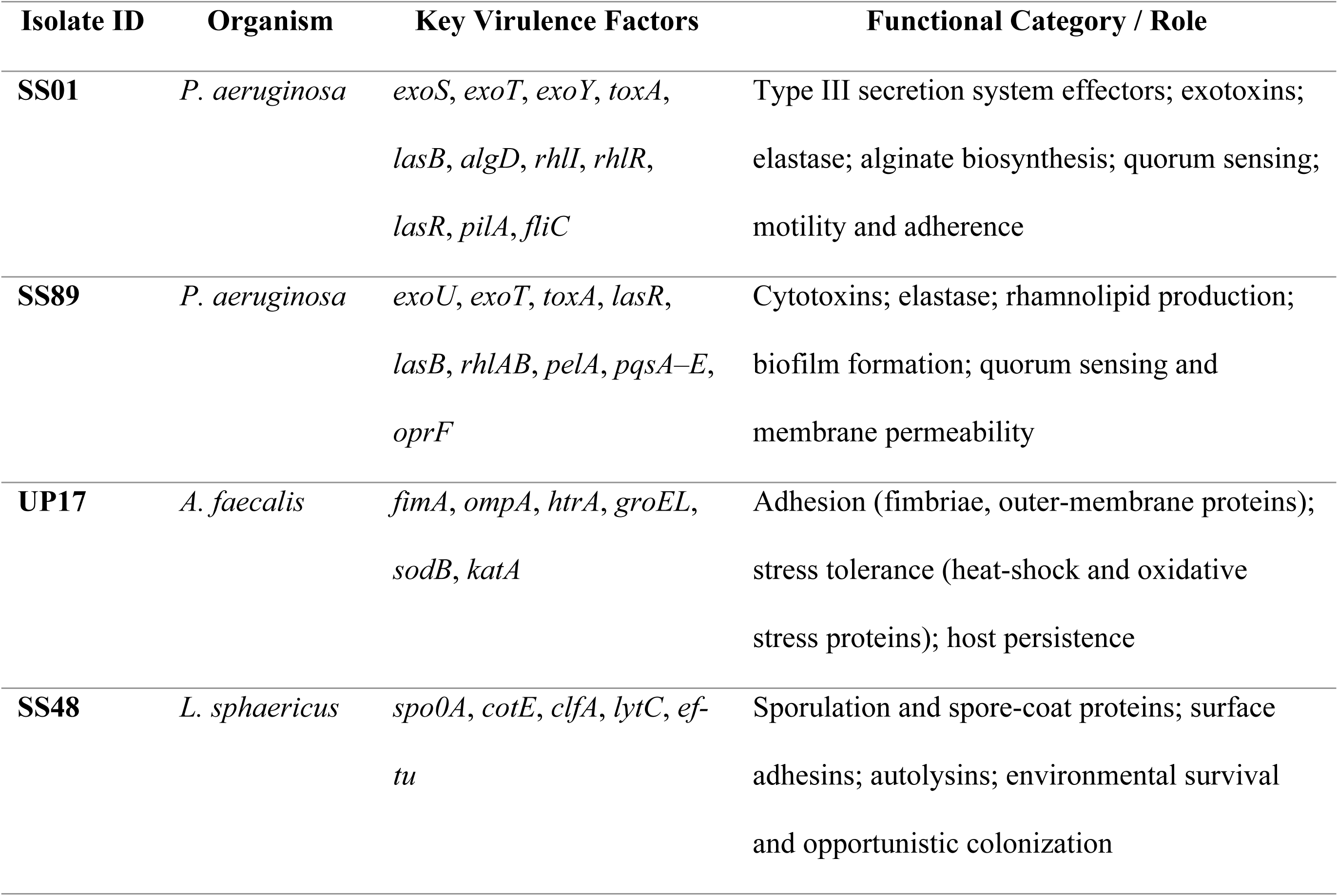
Major virulence factors identified per isolate.

**Table 4:** Summary of genome assembly statistics for the four isolates.

| Isolate ID | Species | Genome size (Mb) | GC content (%) | N50 (bp) | No. of contigs |
| --- | --- | --- | --- | --- | --- |
| SS01 | *P. aeruginosa* | ~6.3 | ~66.0 | 110,000 | 210 |
| SS89 | *P. aeruginosa* | ~6.2 | ~65.9 | 120,500 | 195 |
| UP17 | *A. faecalis* | ~4.2 | ~57.2 | 85,400 | 250 |
| SS48 | *L. sphaericus* | ~4.8 | ~38.6 | 95,600 | 180 |

**Table 5:** Species confirmation and typing results for the four clinical isolates.

| Isolate ID | Species identified | ANI vs reference genome (%) | Closest 16S rRNA match (% identity) | MLST type (if available) |
| --- | --- | --- | --- | --- |
| SS01 | *P. aeruginosa* | >97% | 99.8% with *P. aeruginosa* (NCBI) | Unknown |
| SS89 | *P. aeruginosa* | >97% | 99.7% with *P. aeruginosa* (NCBI) | ST4714 |
| UP17 | *A. faecalis* | >96% | 99.9% with *A. faecalis* (NCBI) | Not applicable |
| SS48 | *L. sphaericus* | >98% | 99.6% with *L. sphaericus* (NCBI) | Not applicable |

### Genome sequencing and assembly

High-throughput Illumina sequencing generated high-quality reads for all isolates. Following trimming and quality control, each isolate was successfully assembled *de novo*. Genome sizes were approximately 6.3 Mb (*P. aeruginosa* isolates) to 4.2 Mb (*A. faecalis*) and 4.8 Mb (*L. sphaericus*), with GC content values consistent with species references. The N50 values indicated good assembly continuity, and the number of contigs was within the expected range for draft bacterial genomes.

### Species confirmation and typing

To ensure accurate taxonomic classification, multiple approaches were applied. Average Nucleotide Identity (ANI) comparisons showed that all four isolates exceeded the 95% threshold typically used to define species boundaries. The *P. aeruginosa* isolates (SS01 and SS89) shared ANI values of >97% with reference *P. aeruginosa* genomes, confirming their identity. Similarly, the UP17 isolate aligned with *Alcaligenes faecalis* reference genomes at >96% ANI, while SS48 demonstrated >98% similarity with *Lysinibacillus sphaericus*, supporting its classification as this species.

Complementary analysis of 16S rRNA gene sequences further validated these assignments. BLASTn searches of the extracted 16S rRNA regions against the NCBI nucleotide database revealed >99% identity with the expected species in each case, in line with current bacterial taxonomy standards. For the *P. aeruginosa* isolates, multilocus sequence typing (MLST) provided finer strain-level resolution. SS01 and SS89 were assigned to distinct sequence types (STs), indicating that despite both being isolated from surgical site infections, they were not clonally related. SS89 was assigned to ST4717 whereas on SS01 no allele hit was detected for the *ppsA* locus during MLST analysis. This suggests independent infection sources or diverse circulating lineages of *P. aeruginosa* within the hospital setting. MLST also enables comparison with international strain collections, which will be useful for tracking the epidemiology of these isolates. Together, the ANI, 16S rRNA BLAST, and MLST results provided strong confirmation of species identity and highlighted strain-level diversity among the Tanzanian *P. aeruginosa* isolates.

### Comparative genomics and phylogenetic placement

Core genome analysis revealed both shared and unique gene sets across the four isolates and their reference genomes. SNP-based phylogenetic trees positioned SS01 and SS89 within distinct *P. aeruginosa* lineages, suggesting independent infection sources. UP17 clustered with *A. faecalis* strains from clinical and environmental backgrounds, highlighting its potential as an emerging uropathogen. SS48 grouped within the *L. sphaericus* clade but remained phylogenetically distinct from most environmental reference strains, suggesting genomic divergence linked to its adaptation to hospital-associated infections. Visualization in iTOL highlighted the evolutionary separation of isolates, while heatmaps showed overlaps in resistance determinants (particularly β-lactamases and efflux systems) across species.

## Discussion

This study explored the genomic and antimicrobial resistance profiles of four clinically significant bacterial isolates obtained from Tanzanian hospitals. The isolates included two *Pseudomonas aeruginosa* (SS01 and SS89) from surgical site infections (SSIs), *Alcaligenes faecalis* (UP17) from a urinary tract infection (UTI) in a pregnant woman, and *Lysinibacillus sphaericus* (SS48) from an SSI. The combined genomic and phenotypic data revealed diverse bacterial species that have independently developed multidrug resistance (MDR), reflecting the growing challenge of antimicrobial resistance (AMR) in hospital settings within low- and middle-income countries.

All four isolates exhibited resistance to commonly used antibiotics, including ciprofloxacin, ceftriaxone, meropenem, amoxicillin–clavulanate, and sulfamethoxazole–trimethoprim. These findings align with the global increase in MDR pathogens, particularly in resource-limited settings where antimicrobial misuse and poor infection control measures drive selective pressure (29,30). The World Health Organization (WHO, 2024) classifies carbapenem-resistant *P. aeruginosa* as a ‘critical priority pathogen,’ underscoring the need for enhanced surveillance and stewardship strategies. The observed resistance to broad-spectrum antibiotics such as carbapenems and fluoroquinolones mirrors previous Tanzanian reports documenting high rates of AMR in *P. aeruginosa* isolated from SSIs and intensive care unit environments (30,32,33). Similar patterns have been reported in Kenya and Uganda, where widespread carbapenemase-producing *P. aeruginosa* and Acinetobacter species have been isolated from hospitalized patients (34). This regional similarity highlights the transboundary spread of AMR genes, often facilitated by horizontal gene transfer and antibiotic selection pressure across healthcare systems.

MLST analysis revealed genetic diversity among the *P. aeruginosa* isolates, with SS01 and SS89 belonging to distinct sequence types ST2317 and ST4714, respectively. Although both were recovered from SSIs, they were genetically unrelated, suggesting multiple sources of infection or colonization within the hospital environment. These findings are consistent with reports that *P. aeruginosa* exhibits extensive genetic diversity and adaptability across hospital and environmental niches (35). Interestingly, SS01 lacked a detectable *ppsA* allele, indicating a possible gap in sequence coverage or allelic divergence not captured in current databases. This limitation emphasizes the need for complementary methods such as core-genome MLST (cgMLST) or single nucleotide polymorphism (SNP)-based phylogenetic analysis for finer resolution.

The *A. faecalis* isolate (UP17) demonstrated an MDR profile consistent with global trends of increasing resistance among opportunistic pathogens. While *A. faecalis* is commonly associated with opportunistic infections, recent studies have shown that it can cause significant morbidity in immunocompromised patients and pregnant women (Huang et al., 2020). Its resistance to β-lactams, aminoglycosides, and tetracyclines complicates treatment and underscores the importance of accurate identification in clinical microbiology laboratories (11,36–38).

On the other hand, *L. sphaericus* (SS48) represents an emerging clinical pathogen traditionally known for its environmental and entomopathogenic roles. However, recent clinical reports, including cases of endocarditis and peritonitis, have implicated this bacterium in opportunistic infections. The presence of multiple AMR genes in SS48 suggests possible acquisition of resistance determinants from environmental reservoirs through horizontal gene transfer. This finding underscores the ‘One Health’ perspective, where environmental microbes may evolve into opportunistic pathogens under selective hospital pressures (13,14,39).

The convergence of AMR determinants across diverse species from different infection types (SSIs and UTIs) suggests shared ecological and clinical drivers. Poor antibiotic stewardship, suboptimal sterilization, and cross-contamination in hospital environments likely facilitate the persistence and spread of MDR organisms (30,40–42). Strengthening infection prevention and control (IPC) programs and routine genomic surveillance could help detect emerging resistant clones early. Moreover, locally generated genomic data can inform empiric treatment guidelines tailored to Tanzanian hospitals rather than relying on external data that may not reflect local resistance patterns.

This study’s main limitation is the small number of isolates analyzed, which limits generalizability. Additionally, short-read sequencing may not fully capture mobile genetic elements such as plasmids or integrons. Future research should incorporate long-read sequencing technologies, longitudinal environmental sampling, and patient linkage data to better understand AMR transmission dynamics. Expanding the genomic library of Tanzanian isolates will also enhance regional representation in global AMR databases, facilitating cross-country comparisons and outbreak tracing. Nevertheless, the findings of this study provide valuable insights into the genomic diversity and AMR mechanisms of *P. aeruginosa*, *A. faecalis*, and *L. sphaericus* isolated from Tanzanian hospitals..

## Conclusion

This study highlights the growing threat of multidrug-resistant bacterial infections in Tanzanian hospitals and the importance of integrating genomic surveillance into clinical practice. The genetic diversity observed among *P. aeruginosa* isolates suggests that infections may arise from different sources rather than a single outbreak strain, while the detection of resistant *A. faecalis* and *L. sphaericus* indicates that both clinical and environmental bacteria contribute to the problem. These findings call for urgent action to strengthen infection prevention, antibiotic stewardship, and genomic monitoring programs in Tanzanian healthcare settings. A coordinated “One Health” approach linking hospitals, laboratories, and environmental surveillance will be essential to slow the spread of antimicrobial resistance and protect the effectiveness of existing treatments.

## Data Availability

Data can be made available upon request to the corresponding author

## Acknowledgments

The authors would like to acknowledge the local (regional and district) medical officers for their administrative support, the General Director and staff of St. Francis Referral hospital, the fish vendors and the patients in the surgical wards for their consent to participate in the study. The authors also extend their gratuity to the laboratory technicians at SFUCHAS and ST. Francis Regional Referral Hospital for their tiresome volunteering during the data processing.

## Notes

### Competing Interest Statement

The authors have declared no competing interest.

### Funding Statement

This study was funded by the Danish Fellowship Centre (DFC) under the Knowledge in Action Grant (Grant ID - 21-EC02-KU).

### Author Declarations

The study was approved by the National Institute for Medical Research (NIMR/HQ/R.8a/Vol.IX/4098, the Local government authority of Ifakara Town Council and the Participant consents

